# Epidemiology of developmental dyslexia: A comparison of DSM-5 and ICD-11 criteria

**DOI:** 10.1101/2020.12.18.20248189

**Authors:** Cécile Di Folco, Ava Guez, Hugo Peyre, Franck Ramus

## Abstract

The two major medical classifications (ICD-11 and DSM-5), define diagnostic criteria for developmental dyslexia that partly differ and that are open to multiple interpretations, inducing different prevalence estimates and discordant cases. The present study evaluates the prevalence of developmental dyslexia for the first time in France in an extensive population representative of French sixth-graders (N=25,000), investigating the consequences of using one classification or the other, and of the different ways of implementing each criterion. Moreover, students diagnosed with dyslexia were compared with the reference population in the other available characteristics. Overall, prevalence estimates ranged from 1.3% to 17.2% depending on the criteria and thresholds used. A reasonable set of criteria and thresholds (−1.5 SD below mean for reading score, -0.5 SD for achievement) yielded a prevalence of dyslexia in France of 6.6% according to DSM-5 and 3.5% according to ICD-11. Factors that had the greatest influence on prevalence estimates were the criteria relative to 1) IQ, and 2) impact on academic achievement. DSM-5, being more liberal than ICD-11 on the IQ criterion, included more cases with relatively low IQ and thus yielded higher prevalence estimates. Compared with the reference population, children with dyslexia were more likely to be boys, to be schooled in a disadvantaged area, and to have lower SES, IQ, and math results. Our results emphasize that the choice of classification and the operationalization of specific criteria have a large impact on who is diagnosed with dyslexia.

## Introduction

Over the last decades, developmental dyslexia has received various names and has been given different definitions. According to the World Health Organization’s (WHO) most recent classification (ICD-11), it is currently known as “developmental learning disorder with impairment in reading” and defined by “significant and persistent difficulties in learning academic skills related to reading, such as word reading accuracy, reading fluency, and reading comprehension”. It markedly affects performance in reading, and is not due to “a disorder of intellectual development, sensory impairment (vision or hearing), neurological disorder, lack of availability of education, lack of proficiency in the language of academic instruction, or psychosocial adversity.” Although it is acknowledged as a real public health issue, the exact prevalence of dyslexia remains a vexed issue, with estimates varying from 2% to 20% across studies.

There are a number of reasons why estimates may vary. One is of course the use of different definitions of dyslexia. Interestingly, the two most widely-used classifications have recently been updated: the APA’s Diagnostic and Statistical Manual in its 5^th^ version (DSM-5) [1], and the International Classification of Diseases in its 11^th^ version (ICD-11) [2]. As we will see below, although the WHO has overall followed the APA quite closely, notably in the grouping of disorders of reading, writing and mathematics, the two classifications crucially differ in certain criteria used in the definition of reading disorder, potentially leading to the identification of different individuals, and to different prevalence estimates. It is therefore important to evaluate the potential discrepancies between the two classifications, and the consequences of using one or the other. We are not aware of any previous study doing so. This will therefore be the main purpose of the present study.

A second reason why prevalence estimates may vary is that dyslexia seems to manifest differently in different writing systems, with greater prevalence in opaque orthographies such as English than in transparent ones such as Italian [3]. French presumably lies in-between [4,5]. Unfortunately, there is a dearth of prevalence studies in France, a gap that this study also aims to fill.

Thirdly, prevalence estimates will depend on the severity threshold chosen for the definition of a “significant” difficulty. Of course, given a certain distribution of reading ability across the population, any cut-off is arbitrary [6,7]. However, prevalence does not just mathematically follow from the chosen threshold, for several reasons that are often under-appreciated. Firstly, the distribution of reading ability is often assumed to be normal, but actual distributions may deviate from normality. Thus, the percentage of cases below a given threshold remains an empirical question, rather than a mathematical deduction. Secondly, prevalence will depend on how exactly the criteria given in official classifications are operationalised. For instance, as emphasised in the ICD-11 definition, reading ability is multifaceted, so prevalence will vary depending on 1) whether one considers reading accuracy, fluency, and/or comprehension, on 2) whether one uses one, two or three reading measures, and on 3) whether one applies the threshold to each measure separately (using an OR or an AND connector), or to a composite of the different measures. Prevalence may also vary depending on whether an absolute threshold is applied to the distribution of reading ability, or whether the threshold on reading ability is applied relative to the level of intellectual functioning. This issue of whether dyslexia should be defined according to a discrepancy or not is highly controversial[8], and this is one important criterion on which DSM-5 and ICD-11 crucially differ. Finally, definitions of dyslexia are not exclusively based on reading ability, but also include a number of exclusion criteria, which contribute to reducing the prevalence compared to the raw number of individuals with a significant difficulty in reading.

The largest epidemiological studies were conducted in English-speaking countries. In the United-States, the highest prevalence estimate is given by Shaywitz, Fletcher and Shaywitz, (1994), who evaluated that 17.5% of students were dyslexic, according to a low-achievement criterion only (reading score inferior to -0.67 SD; N=445). Katusic, Colligan, Barbaresi, Schaid and Jacobsen (2001) used four different methods on a sample of 5,718 children from a birth cohort and obtained prevalences ranging from 5.3% (with a discrepancy criterion) to 11.8% (with a low-achievement criterion). Lindgren, De Renzi and Richman (1985) compared three definitions on American and Italian students samples, and found prevalences varying between 4.5% to 12% in the United-States and 3.6% to 8.5% in Italy, the difference being attributed to differences in orthographic transparency. In the United Kingdom, Yule, Rutter, Berger and Thompson (1974) found 3.61% of dyslexics in the Isle of Wight (N=1,134) and 9.26% in London (N=1,643), whereas Rodgers (1983) found a prevalence of 2.3% (N=8,836) in a sample representative of the national population. They used the same definition of dyslexia, the children were the same age (10 years old), the test was not exactly the same but involved reading comprehension in both cases.

In France, however, no such research has been done. The results found in English-speaking countries cannot be generalized to French, whose orthography is slightly more transparent than that of English, leading to the expectation of a lower prevalence of dyslexia. A few prevalence estimates can be extracted from studies that did not primarily aim at calculating it. These studies defined dyslexia as under-achievement, without any discrepancy criterion. Estimates include 12% (N=199 7-year-old children) according to Plaza et al. (2002), and 7.5% (N=485 8-year-old children) according to Zorman, Lequette and Pouget (2004), who added a phonological deficit criterion. The limited size and non-representativeness of the samples make any generalization to the French population impossible.

Beyond prevalence, there remain many controversial issues, such as the sex-ratio in dyslexia, the role of socioeconomic factors, and the potential role of left-handedness, which may in turn depend on the specific choice of diagnostic criteria.

Thus, the aims of the present study are to 1) estimate the prevalence of dyslexia in France, using both DSM-5 and ICD-11 criteria, comparing them for the first time and evaluating their concordance; 2) evaluate the impact of each diagnostic criterion on prevalence estimates; 3) characterise the population of dyslexic individuals, and how this varies according to diagnostic criteria. We do so on a large representative sample of French 6^th^ grade pupils.

## Methods

### Participants

The *Direction de l’Evaluation, de la Prospective et de la Performance* (DEPP), French Ministry of Education, conducted a large prospective study following students from the beginning (grade 6) to the end of French middle school (grade 9). The National Council for Statistical Information (CNIS) approved this study, ensuring public interest and conformity with ethical, statistical and confidentiality standards. The sample consisted of about 35,000 randomly selected students among the 760,000 entering middle school in 2007, with a higher sampling rate for schools in disadvantaged areas. A more detailed description of the study can be found in [15].

For the current purpose, we restricted the analysis to Grade 6, a stage at which the prevalence of dyslexia stabilises according to Katusic, Colligan, Barbaresi, Schaid and Jacobsen (2001)’s longitudinal follow-up. We excluded students who had missing values for intelligence and reading scores in grade 6, as well as academic performance score in grade 6. Given an unexpectedly high number of students scoring 0 at the intelligence test, suggesting either a refusal to take the test, low engagement in the task, or problems with administration and scoring, we also excluded those students. Children who repeated a year were not excluded. This process is summed up in the flowchart (Supplementary Figure S1). 25,041 students were thus included in the present study. Included and excluded participants differed in observed characteristics (see Supplementary Table S1).

### Measures

The DEPP conducted standardised evaluations in French, mathematics, non-verbal intelligence and perceived self-efficacy, in paper/pencil format. These tests were administered collectively, supervised by teachers. Children’s socio-economic status was estimated based on a questionnaire filled by parents (or legal guardians). The following variables were used for the present analysis.

### Tests

Examples of test items can be found in supplementary methods.

#### Reading comprehension

Students were asked to read a short text, then answer 5 open-ended comprehension questions. They did so for 3 different texts, in a maximum of 12 minutes (total: 15 items). Examples of test items for this and other tests can be found in Supplementary Methods.

#### Phonological awareness

Participants were given a list of 5 written words and had to tick the one that did not share a common sound with the others. There were 10 trials.

#### Grammar

Students had to fill in the blanks in 3 short texts with logical connectors, determiners or pronouns (20 items, open-ended questions).

#### Mathematics

Students had to answer 48 questions (26 open and 22 multiple choice) involving problem solving, logic, mental arithmetic, notions of time and units.

#### Nonverbal intelligence

The *Raisonnement sur Cartes de Chartier* test (Chartier’s Reasoning Test on Playing Cards) assesses logical reasoning skills using playing cards [16,17]. Inspired from Raven’s progressive matrices, it consists of 30 items in which children must find the missing card (from a deck of 4 suits of 10 cards) in an array composed of 4 to 12 cards, within a time limit of 20 minutes.

##### Academic performance

At the beginning of grade 6 (entrance in middle school), all French students have to take national assessments in French and Mathematics. These tests are administered in schools by the school teachers and have no stakes for the students: they are only meant to give teachers and parents an idea of the students’ levels. We took the mean of results in French and Mathematics as an indicator of academic performance in grade 6.

##### Perceived self-efficacy

In grade 6, students answered questions from the Children’s Perceived Self-Efficacy scales [18], closely translated into French. It is a 37-item questionnaire from which factors representing perceived academic self-efficacy, social self-efficacy and self-regulatory efficacy were extracted. For each item, students had to evaluate their ability to perform a given activity using a 5-point Likert scale. The perceived academic self-efficacy score measures students’ perceived ability to manage their learning, to master different academic subjects (mathematics, science, etc…), and to fulfill parents’ and teachers’ expectations. The perceived social self-efficacy score measures efficacy regarding leisure group activities, the ability to form and maintain social relationships and manage interpersonal conflicts, and self-assertiveness. Lastly, the perceived self-regulatory efficacy score measures students’ perceived ability to resist peer pressure to engage in high-risk activities (alcohol, drugs, transgressive behaviors).

##### Motivation

Students’ academic motivation was assessed in grade 6 with questions derived from the Academic Self-Regulation Questionnaire (SRQ-A) [19], adapted and translated into French [20]. This is a self-report measurement which assesses individual differences in motivational styles. The items asked students the reasons why they do their home and class work, and try to do well in school. Each item provides a possible reason that represents a certain motivational style (for example: “I do my classwork because I want to learn new things”, or “I do my classwork because I’d be ashamed of myself if it didn’t get done”). Each item was answered using a 5-point Likert scale. Three factors were extracted: intrinsic motivation, extrinsic motivation, and amotivation.

### Variables from the family questionnaire

#### Handedness

Parents reported whether the child was right-handed (N=20,466), left-handed (N=2,977) or ambidextrous (N=300). We grouped together left-handed and ambidextrous; our handedness variable is thus binary (right-handed versus non-right-handed).

#### Whom the child lives with

Parents or legal guardian reported whether the child lived with his family (several options) or in child care.

#### Serious disease

Parents were asked to indicate if the child had any serious illness, and specify the age range of onset of the disease (before age 1, between 1 and 5, between 6 and 9, between 10 and 13). No details regarding the duration or type of the disease were reported.

#### Year of arrival in France

In case the child was born abroad, the year of arrival in France was reported amongst: between 1993 and 1995, between 1996 and 1998, between 1999 and 2001, between 2002 and 2004, between 2005 and 2007.

#### Household monthly income

Parents reported the household monthly income. Given the skewed distribution, we used the natural logarithm of income.

#### Parental education

Parents’ highest diploma was converted into years of education for each parent (from 0 to 18.5 years –18.5 years corresponding to a graduate degree). Parental education was then estimated as the mean of the two parents’ education (when one was missing, only the other one was used).

*Socio-economic status* (SES) was computed as the mean of z-scored household income and parental education (when one was missing, only the other one was used).

#### Disadvantaged school

Obtained from the ministry’s central database, this variable indicated whether the child’s school belonged to either *Réseau de Réussite Scolaire* or *Réseau Ambition Réussite*, two networks of disadvantaged areas targeted for priority education policy.

### Analysis of diagnostic criteria

Table 1 lists each of the diagnostic criteria included in ICD-11’s definition of “developmental learning disorder with impairment in reading” and in DSM-5’s definition of “specific learning disorder, with impairment in reading”. While the wording differs, many of the criteria are similar, and are therefore placed on the same line in the table for comparison. In the third column, we indicate whether and how we took each criterion into account in the present analysis.

**Table 1.** Systematic comparison of ICD-11 and DSM-5 criteria for developmental dyslexia, and their operationalisation in the present study.

| ICD-11 criteria | DSM-5 criteria | Operationalisation | Comment |
| --- | --- | --- | --- |
| Persistent difficulties | Presence of at least one of the following symptoms that have persisted for at least 6 months.<br>The learning difficulties begin during school-age years | Application in 6<sup>th</sup> grade meets both persistence and school-age criteria. |  |
|  | despite the provision of interventions that target those difficulties | Not taken into account | No data available. |
| in learning academic skills related to reading, such as word reading accuracy, reading fluency, and reading comprehension. | With impairment in reading:<br>Word reading accuracy<br>Reading rate or fluency<br>Reading comprehension | Score below threshold on reading comprehension measure | No other reading measure available. |
| The individual's performance in reading is markedly below what would be expected for chronological age | The affected academic skills are substantially and quantifiably below those expected for the individual's chronological age, | Reading score x SD below the mean of students born in 1996 and in 6<sup>th</sup> grade in 2007 | x=1, 1.25, 1.5 (default), 1.75, 2 |
| and level of intellectual functioning |  | Reading score x SD below that predicted by linear regression of reading on non-verbal intelligence. | x=1, 1.25, 1.5 (default), 1.75, 2 |
| and results in significant impairment in the individual's academic or occupational functioning. | and cause significant interference with academic or occupational performance, or with activities of daily living | Score at 6<sup>th</sup> grade national evaluations $y$ SD below mean | $y=0, 0.5$ (default), 1 |
| Developmental learning disorder with impairment in reading is not due to a disorder of intellectual development, | The learning difficulties are not better accounted for by intellectual disabilities, | Score above -2SD on non-verbal IQ |  |
| sensory impairment (vision or hearing), | uncorrected visual or auditory acuity, | Not taken into account | No data available. |
| neurological disorder, | other mental or neurological disorders, | Not taken into account | No data available. |
| lack of availability of education, | or inadequate educational instruction. | Exclusion of pupils with "serious illness" before age 9. | No information available on "adequacy" of instruction, or on other causes of school drop-out |
| lack of proficiency in the language of academic instruction, | lack of proficiency in the language of academic instruction, | Exclusion of pupils arrived in France after age 6. | No direct information available on French language proficiency |
| or psychosocial adversity. | psychosocial adversity, | Exclusion of pupils in care. | No information available on other indicators of psychosocial adversity |

In summary, criteria shared by the two classifications include 1) significance/severity of the disorder; 2) school-age manifestation and persistence; 3) specification of relevant reading skills; reference to a norm for chronological age; 5) interference with academic or occupational performance; 6) exclusion of intellectual disability, sensory and neurological disorders, inadequate education, non-proficiency in instruction language, and psychosocial adversity. Criteria where the classifications differ are 1) only the DSM-5 includes an “insufficient response to intervention” criterion; 2) only the ICD-11 includes an IQ-discrepancy criterion.

Some criteria deserve further comment.

- Regarding the IQ-discrepancy threshold (ICD-11), we investigated 1, 1.25, 1.5, 1.75, 2 SD below the reading score linearly predicted by non-verbal IQ. Because the variance of reading scores decreases as non-verbal intelligence increases (a Breusch-Pagan test confirmed heteroscedasticity: *p*<0.0001), we used weighted least squares to estimate the variance of residuals as a function of non-verbal intelligence score (assuming that the variance varies linearly with IQ scores) and to obtain efficient regression estimates (further details can be found in Supplementary Methods).
- The “interference with academic or occupational performance” is a ubiquitous criterion in medical classifications, but is never applied in research and only partly so in clinical settings (beyond the fact that consultation implies some degree of perceived interference). Here, we applied this criterion, using a threshold on the mean score of 6^th^ grade national evaluations (including both French and Mathematics), to represent interference with academic achievement. We investigated thresholds 0, 0.5 and 1 SD below the mean, with 0.5 SD as the default threshold, reasoning that “interference” does not imply as stringent thresholds as “disorder”.

### Statistical analyses

Prevalence of dyslexia was calculated using the full set of criteria and each threshold for both ICD-11 and DSM-5. The impact of the various criteria on prevalence was then investigated by:

- Varying the severity threshold (from 1 to 2 SD below mean). For ICD-11, the IQ-discrepancy threshold was kept equal to the severity threshold, for consistency.
- Varying the interference threshold (from 0 to 1 SD below mean).
- Relaxing each of the other criteria one at a time (using the 0.5 SD default threshold for interference).

Agreement between ICD-11 and DSM-5 diagnoses was compared using a contingency table (using default thresholds).

The dyslexic and control populations were then compared on all the available variables: reading comprehension, grammar, phonology, mathematics, non-verbal intelligence, mean score at the national evaluations, self-efficacy, motivation, SES, schooling in priority education area, sex, handedness, and grade repetition, using weighted t-tests and chi-square tests.

Finally, given the interest in “twice exceptional children” who show both high IQ (>130) and a learning disability, and given uncertainties on their prevalence [21–24], we also estimated the prevalence of dyslexia separately in high-IQ and in other pupils.

All analyses were conducted in R (survey, questionr and weights packages), using weights based on an exhaustive baseline survey, in order to adjust the results to a representative sample of the population, and non-response propensity weights to account for selective inclusion (further details can be found in Supplementary Methods and Table S2).

## Results

Descriptive statistics for the included participants and comparison with the excluded participants are provided in Table S1.

### Impact of classification, thresholds and criteria on dyslexia prevalence

Table 2 presents the prevalence obtained using the two classifications, depending on severity thresholds, and academic achievement threshold. Furthermore, we investigated the impact of each of the other criteria by reporting the prevalence when this criterion is relaxed (at the default achievement threshold -0.5 SD). For instance, according to ICD-11, when using a -1.5 SD threshold on reading ability and a -0.5 SD threshold on academic achievement, the prevalence of dyslexia is 3.5%. When relaxing the IQ criterion (IQ>70 and reading < -1.5 SD score predicted by IQ), the prevalence raises to 7.9%. Using the same thresholds, the prevalence of dyslexia according to DSM-5 is 6.6%. When relaxing the IQ criterion (IQ>70), the prevalence raises to 7.9%, just like according to ICD-11, since the two classifications differ only in the IQ criterion in the present study (the other difference, absence of response to intervention, could not be taken into account). Using the same thresholds, when the academic achievement criterion is relaxed, prevalence raises from 3.5 to 5.1% according to ICD-11 and from 6.6 to 9.3% according to DSM-5. All the other criteria (as implemented here) have a much lower impact on prevalence.

**Table 2.** Prevalence of dyslexia according to ICD-11 and DSM-5, at different thresholds, using all criteria and relaxing one criterion at a time. Default thresholds are in bold. Source : MENESR DEPP, Panel 2007.

| ICD-11 |  |  |  |  |  |  |  |  |
| --- | --- | --- | --- | --- | --- | --- | --- | --- |
| Severity<br>threshold | All criteria |  |  | Relaxing 1 criterion |  |  |  |  |
|  | Achievement threshold |  |  | IQ (absolute<br>threshold +<br>discrepancy) | Academic<br>achievement | Serious<br>illness | Language<br>proficiency | Psychosocial<br>adversity |
|  | 0 SD | **-0.5**<br>**SD** | -1 SD |  |  |  |  |  |
| -1 SD | 9.6 | **7.3** | 4.7 | 15.0 | 12.3 | 7.6 | 7.7 | 7.4 |
| -1.25 SD | 6.4 | **5.1** | 3.5 | 11.2 | 8.0 | 5.3 | 5.4 | 5.2 |
| **-1.5 SD** | **4.3** | **3.5** | **2.5** | **7.9** | **5.1** | **3.7** | **3.7** | **3.6** |
| -1.75 SD | 2.6 | **2.2** | 1.7 | 5.1 | 3.0 | 2.3 | 2.4 | 2.2 |
| -2 SD | 2.0 | **1.7** | 1.3 | 5.1 | 2.3 | 1.8 | 1.8 | 1.7 |
| DSM-5 |  |  |  |  |  |  |  |  |
| Severity<br>threshold | All criteria |  |  | Relaxing 1 criterion |  |  |  |  |
|  | Achievement threshold |  |  | IQ (absolute<br>threshold) | Academic<br>achievement | Serious<br>illness | Language<br>proficiency | Psychosocial<br>adversity |
|  | 0 SD | **-0.5**<br>**SD** | -1 SD |  |  |  |  |  |
| -1 SD | 17.2 | **13.1** | 8.4 | 15.0 | 21.4 | 13.6 | 13.7 | 13.3 |
| -1.25 SD | 12.0 | **9.7** | 6.6 | 11.2 | 14.3 | 10.0 | 10.1 | 9.8 |
| **-1.5 SD** | **7.9** | **6.6** | **4.8** | **7.9** | **9.3** | **6.9** | **7.0** | **6.7** |
| -1.75 SD | 4.8 | **4.2** | 3.3 | 5.1 | 5.4 | 4.3 | 4.4 | 4.2 |
| -2 SD<sup>a</sup> | 4.8 | **4.2** | 3.3 | 5.1 | 5.4 | 4.3 | 4.4 | 4.2 |

<sup>a</sup> The last 2 lines showing identical numbers is due to the discreteness of reading comprehension scores, and the absence of any cases between -1.75 SD and -2 SD.

Figure 1 shows the relationship between reading comprehension and non-verbal IQ scores across the entire population, with lines showing both severity and IQ thresholds according to each classification. It illustrates the main difference between the two classifications: In ICD-11, dyslexic individuals are in the bottom right trapezium, while in DSM-5 they are in the bottom right rectangle. Individuals in the yellow triangle in Figure 1C are those diagnosed as dyslexic by DSM-5 but not by ICD-11, because their reading score is not sufficiently discrepant with their IQ.

**Figure 1.**
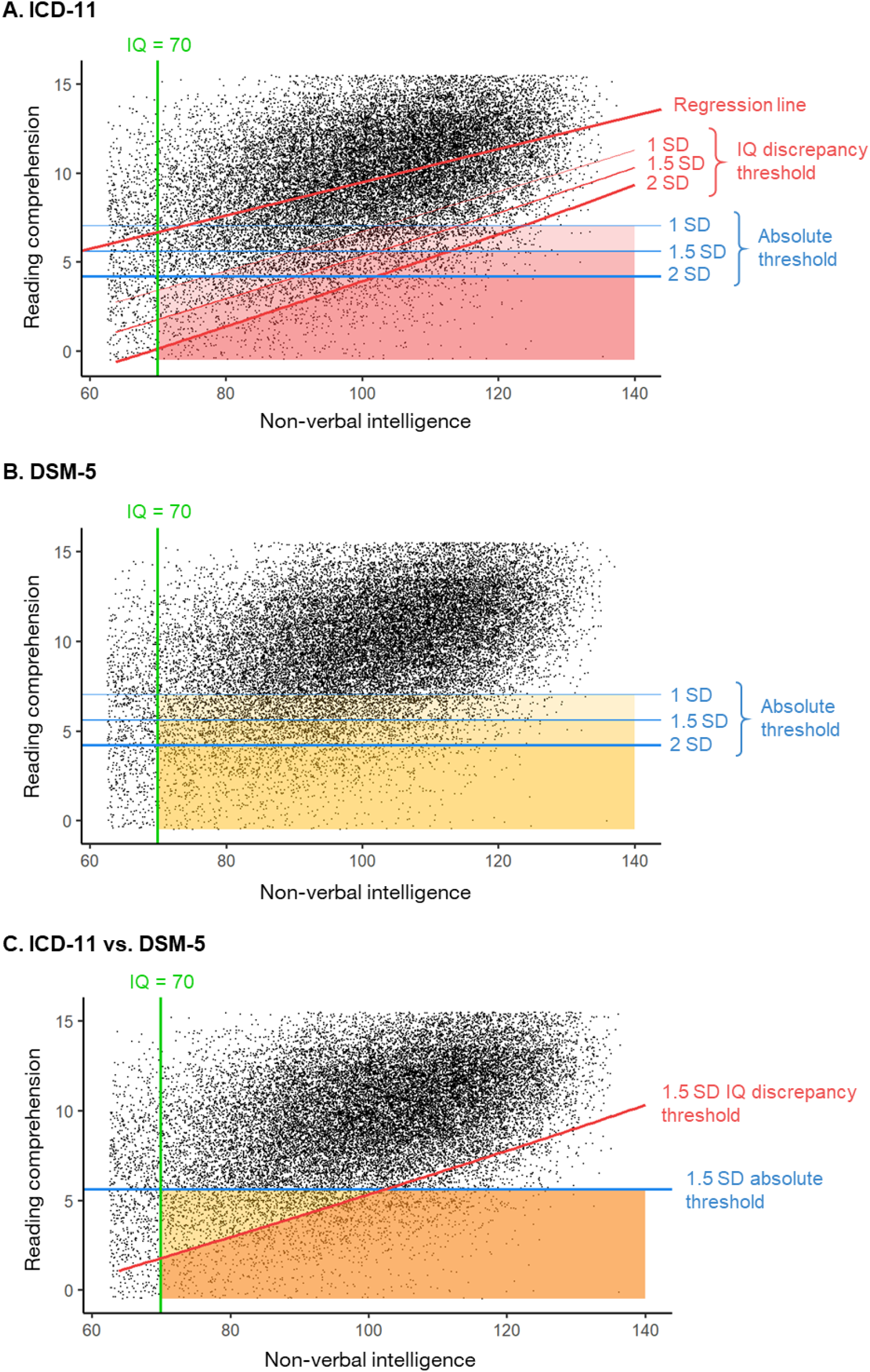
Reading comprehension as a function of non-verbal intelligence in the entire population. Lines represent different severity thresholds and their application by ICD-11 and DSM-5. A) ICD-11. B) DSM-5. C) Comparison of ICD-11 and DSM-5 criteria. Regions in shades of red represent individuals diagnosed by ICD-11. Regions in shades of yellow represent individuals diagnosed by DSM-5. The region in orange (C) represents individuals diagnosed by both classifications. Scores on both dimensions are jittered for better visibility (hence some seemingly negative reading scores). Source : MENESR DEPP, Panel 2007.

### Concordance between the two classifications

Table 3 shows the contingency table of the classification into dyslexic and normal reader according to both classifications. Using the default thresholds (−1.5 SD for reading severity and

**Table 3.** Contingency table for dyslexic and normal readers according to DSM-5 and ICD-11. Source : MENESR DEPP, Panel 2007.

| DSM-5 |  |  |  |  |
| --- | --- | --- | --- | --- |
| ICD-11 | Dyslexic | Normal reader | *Total* | % |
| Dyslexic | 884 | 0 | 884 | 3.53 |
| Normal reader | 778 | 23379 | 24157 | 96.47 |
| *Total* | 1662 | 23379 | 25041 | 100 |
| % | 6.64 | 93.36 | 100 |  |

-0.5 SD for achievement), DSM-5 diagnoses almost twice as many individuals as ICD-11 (884 for ICD-11, 1662 for DSM-5). All the individuals diagnosed by ICD-11 are also diagnosed by DSM-5 (in the absence of the response to intervention criterion), but DSM-5 additionally includes a large number of individuals with a reading score above the reading-IQ discrepancy threshold (778 individuals).

### Characteristics of individuals diagnosed as dyslexic

Table 4 reports the results on all the available variables for individuals diagnosed or not with dyslexia, according to ICD-11 and DSM-5. By definition, dyslexic readers scored much lower than normal readers in reading comprehension (Cohen’s d≈-2.8), as well as in other measures of French language (grammar: d≈-1.5; phonology: d≈-0.9), in Mathematics (d≈-1.4), and in overall academic achievement (d≈-1.9). On average, they had a lower SES (d≈-0.7), they were twice as likely to be in a priority education area, 3.5 times as likely to have repeated a grade, 70% more likely to be a boy, and 4 to 25% more likely to be non-right-handed. They also showed lower ratings in various measures of self-efficacy and motivation. In all these measures, differences between the two groups were generally similar between the two classifications. Only IQ showed a substantial difference: dyslexic readers had on average a lower IQ than normal readers, by d=-0.44 according to ICD-11 and d=-0.91 according to DSM-5.

**Table 4.** Descriptive Statistics for individuals diagnosed by ICD-11, DSM-5 and normal readers, computed using the -1.5 SD severity criterion, the - 0.5 SD achievement criterion, and weighted data. Source : MENESR DEPP, Panel 2007.

| Outcome variable | ICD-11 |  |  |  |  |  | DSM-5 |  |  |  |  |  |
| --- | --- | --- | --- | --- | --- | --- | --- | --- | --- | --- | --- | --- |
|  | Dyslexic |  | Normal reader |  | Difference |  | Dyslexic |  | Normal reader |  | Difference |  |
|  | N=884 |  | N=24,157 |  | (Dyslexic -<br>Normal reader) |  | N=1,662 |  | N=23,379 |  | (Dyslexics -<br>Others) |  |
|  |  |  |  |  | *d or* |  |  |  |  |  | *d or* |  |
|  | *M* | *SD* | *M* | *SD* | *Odds* | *p* | *M* | *SD* | *M* | *SD* | *Odds* | *p* |
|  | (or %) |  | (or %) |  | Ratio |  | (or %) |  | (or %) |  | Ratio |  |
| *Reading Scores* |  |  |  |  |  |  |  |  |  |  |  |  |
| Reading comprehension | 2.84 | 1.54 | 9.72 | 2.89 | -2.96 | <.0001 | 3.6 | 1.47 | 9.89 | 2.77 | -2.84 | <.0001 |
| Grammar | 3.4 | 2.56 | 8.8 | 4.25 | -1.54 | <.0001 | 3.62 | 2.55 | 8.97 | 4.19 | -1.54 | <.0001 |
| Phonology | 4.79 | 2.27 | 6.81 | 2.15 | -0.91 | <.0001 | 4.89 | 2.25 | 6.87 | 2.12 | -0.9 | <.0001 |
| Mathematics | 16.24 | 5.98 | 26.33 | 8.71 | -1.35 | <.0001 | 15.83 | 5.58 | 26.7 | 8.57 | -1.5 | <.0001 |
| National assessment score | 33.6 | 10.84 | 60.27 | 16.81 | -1.89 | <.0001 | 34.2 | 10.23 | 61.11 | 16.33 | -1.97 | <.0001 |
| Intelligence score (non-verbal) | 94.28 | 11.76 | 100.21 | 15.06 | -0.44 | <.0001 | 88.64 | 11.62 | 100.81 | 13.89 | -0.91 | <.0001 |
| Socio-economic index | -0.61 | 0.88 | 0.02 | 1 | -0.67 | <.0001 | -0.6 | 0.88 | 0.04 | 0.99 | -0.68 | <.0001 |
| Priority Education (% , OR) | 27.1 |  | 15.2 |  | 2.07 | <.0001 | 28.2 |  | 14.8 |  | 2.27 | <.0001 |
| Repeater (% , OR) | 54.1 |  | 15.8 |  | 3.28 | <.0001 | 52.4 |  | 14.6 |  | 3.81 | <.0001 |
| Boys (% , OR) | 64.6 |  | 50.5 |  | 1.79 | <.0001 | 61.2 |  | 50.3 |  | 1.56 | <.0001 |
| Non-right-handed (% , OR) | 14.2 |  | 13.8 |  | 1.04 | 0.7505 | 16.4 |  | 13.6 |  | 1.25 | 0.00293 |
| *Perceived self-efficacy* |  |  |  |  |  |  |  |  |  |  |  |  |
| Self-regulation | -0.83 | 1.09 | 0.04 | 0.97 | -0.84 | <.0001 | -0.79 | 1.11 | 0.06 | 0.96 | -0.9 | <.0001 |
| Social self-efficacy | -0.29 | 1.17 | 0.01 | 0.98 | -0.28 | <.0001 | -0.26 | 1.15 | 0.01 | 0.98 | -0.26 | <.0001 |
| Academic self-efficacy | -0.66 | 1.07 | 0.01 | 0.98 | -0.65 | <.0001 | -0.6 | 1.08 | 0.03 | 0.97 | -0.62 | <.0001 |
| *Motivation* |  |  |  |  |  |  |  |  |  |  |  |  |
| Intrinsic motivation | -0.11 | 0.98 | -0.01 | 0.92 | -0.1 | 0.0076 | -0.08 | 0.97 | -0.01 | 0.92 | -0.08 | 0.0036 |
| Extrinsic motivation | 0.01 | 0.9 | -0.01 | 0.82 | 0.02 | 0.6489 | 0.042 | 0.88 | -0.01 | 0.82 | 0.06 | 0.032 |
| Amotivation | 0.43 | 1.12 | -0.02 | 0.86 | 0.45 | <.0001 | 0.39 | 1.14 | -0.03 | 0.85 | 0.42 | <.0001 |

Table S3 reports a more detailed analysis of how the sex-ratio varied as a function of each diagnostic criterion. Whereas the sex-ratio (M/F) in the entire population was 1.04, it increased to 1.6 for poor readers (reading score below -1.5 SD). Furthermore, the more severe the threshold, the higher the sex-ratio, from 1.47 at -1 SD to 1.76 at -2 SD. Most exclusion criteria did not affect sex-ratio, however the exclusion of individuals with intellectual disability decreased the sex ratio slightly to 1.54 (at -1.5 SD), suggesting that boys were slightly overrepresented among the very low IQ poor readers. Overall, the sex-ratio for DSM-5 criteria remained close to that for poor readers. However, the addition of the IQ-discrepancy (ICD-11 criteria) significantly increased the sex-ratio, to 1.53 at -1 SD and to 2.22 at -2 SD, suggesting that girls were overrepresented among the non-discrepant poor readers.

Table S4 provides a more detailed analysis of handedness as a function of diagnostic criteria. Overall, non-right-handedness was slightly more prevalent in poor readers (15.7% at -1.5 SD) and in DSM-5 dyslexic readers (16.4%) than in the entire population (13.8%). Prevalence of non-right-handedness increased with the severity threshold (from 15% at -1 SD to 16.5% at -2 SD). However, the application of the IQ discrepancy criterion lowered the prevalence of non-right-handedness (to about 15%), making it non-significantly different from the base rate.

We then considered whether this increased prevalence of non-right-handedness in dyslexia might be due to the sex-ratio in favour of boys, or to the lower IQ. Indeed, we found non-right-handedness in 15.6% of boys and 12.0% of girls. Furthermore, we found that non-righthanders scored on average 2 IQ points lower than right-handers. In order to eliminate biases due to sex and IQ, we ran a logistic regression with diagnostic category as dependent variable, and handedness, sex, IQ and their interactions as independent variables. We found significant effects of IQ, of sex (in DSM-5 only), and of the interaction between sex and IQ on diagnostic category. However, there was no significant effect of handedness, nor any interaction involving handedness, suggesting that the prevalence of handedness does not differ between dyslexic and normal readers, once sex and IQ are controlled.

Finally, we found a prevalence of dyslexia of 0.4% [0.05; 3.0] in high IQ students (nonverbal intelligence > 130) with both DSM-5 and ICD-11 criteria, compared to 3.7% [3.4; 4.0] (ICD-11) and 6.9% [6.6; 7.0] (DSM-5) in the non-high IQ population.

## Discussion

We evaluated the prevalence of dyslexia in France in a large representative population of 6^th^ grade pupils, according to the two widely-used international classifications, and at five different severity thresholds. Prevalence estimates range from 1.3% (ICD-11, -2 SD severity threshold, - 1 SD achievement threshold) to 17.2% (DSM-5, -1 SD severity threshold, 0 SD achievement threshold). Using reasonable compromise thresholds (−1.5 SD for severity, -0.5 SD for achievement), prevalence estimates are 3.5% with ICD-11 and 6.6% with DSM-5. These numbers are within the range described in the literature, although no previous study to our knowledge used exactly the same set of criteria approximating most closely the definitions in DSM-5 and ICD-11. Our estimates are also consistent with the idea that the prevalence of dyslexia in France should be intermediate between that in English-speaking countries (e.g., 7.3% in the USA according to [3]) and that in countries with a more transparent orthography (e.g., 3.6% in Italy according to [3]), although again direct comparison is not possible. Beyond the numbers that are to some extent arbitrary, the present study’s most important contribution lies in the investigation of the consequences of each diagnostic criterion on prevalence estimates and on the characteristics of the population diagnosed with dyslexia.

### Impact of each criterion on prevalence

#### IQ-reading discrepancy

We found that DSM-5 yielded systematically higher prevalence rates than ICD-11, and this is directly attributable to the one criterion that differed between the two: the IQ-reading discrepancy threshold, required by ICD-11 but not by DSM-5. Compared with the DSM-5, applying this threshold additionally excluded from the ICD-11 diagnosis all individuals who had an IQ above 70, and who had a reading score below the severity threshold, but above the discrepancy threshold (illustrated by the region in yellow in Figure 1C). Depending on the specific thresholds, this had the effect of excluding up to half of all individuals diagnosed with dyslexia by DSM-5. Whether or not one implements such a discrepancy criterion has therefore important consequences for many individuals whose reading ability is not substantially below that predicted by their non-verbal intelligence. However, it should also be noted that our reading score being based on reading comprehension must have inflated the correlation between reading and IQ (compared to a reading measure based on reading accuracy and/or fluency), thus may overestimate the impact of the IQ-reading discrepancy criterion.

As a side remark, it may seem paradoxical that the DSM-5 chose to emphasize specificity in the name “specific learning disorder” while simultaneously shunning the IQ-discrepancy criterion that operationalised cognitive specificity, when the ICD-11 preserved this specificity criterion, without emphasizing it in the name of the disorder (“developmental learning disorder”).

#### Insufficient response to intervention

Our finding of a systematically higher prevalence in DSM-5 should be taken with caution, given that we were unable to evaluate the impact of the “insufficient response to intervention” criterion, which is required in DSM-5 only, and which should logically decrease the reported prevalence for DSM-5. We are not aware of any previous study implementing this diagnostic criterion. Therefore, how much DSM-5 prevalence would decrease if the diagnostic criteria were fully implemented is a matter of speculation.

#### Interference with academic performance

Together with IQ-reading discrepancy, this was the other criterion that had a major impact on prevalence estimates. In ICD-11, prevalence ranged from 2.5% with a -1 SD threshold on achievement to 5.1% without such a threshold. In DSM-5, prevalence similarly ranged from 4.8% to 9.3%. Thus again, whether one applies such a criterion has a major impact on the individuals who will or will not receive a diagnosis. The individuals concerned are those who manage, in one way or in another, to reach at least normal academic performance despite their reading difficulties. Whereas medical classifications unanimously consider that there can be no diagnosis without sufficient evidence of impact on one’s functioning, many people consider that even normal and over-achievers may deserve a diagnosis of dyslexia and require help.

#### Exclusion criteria

The exclusion criteria that we were able to partly apply were intellectual disability, lack of education, lack of proficiency in the language of instruction and psychosocial adversity. The intellectual disability criterion excluded between 1 and 2% of children from a diagnosis of dyslexia. All other exclusion criteria had a small impact (at most 0.6%). The exclusion criteria that we were not able to apply were sensory impairment (auditory or visual), neurological disorder, and inadequate instruction (DSM-5). Applying the first two might decrease prevalence by at most another 1%, at least in countries like France where such disorders are infrequent. Arguably, excluding cases with inadequate instruction might have a larger impact, depending on how “adequate instruction” is defined (e.g., more or less strict phonics methods) and implemented in the country of interest.

### Characteristics of dyslexic pupils according to ICD-11 and DSM-5

#### Non-verbal intelligence

Pupils diagnosed with dyslexia had on average slightly lower non-verbal IQ than the rest of the population, and this was more pronounced according to DSM-5 (d=-0.91) than according to ICD-11 (d=-0.44). This difference follows from the reading-IQ discrepancy criterion, whereby ICD-11 excluded a number of pupils with relatively low-average IQ that were included by DSM-5 (Stuebing et al., 2002, found similar results). This lower IQ contrasts with the widespread stereotype of the “normal or superiorly intelligent” dyslexic child, but is a logical consequence of the correlation between general intelligence and reading ability. It is consistent with some previous population-based studies [10] but not others [25]. In the present study, the IQ difference may have been amplified by the application of the “interference with academic performance” criterion, which was not used by most previous studies.

#### Academic achievement

By definition, dyslexic readers scored much lower than normal readers in reading comprehension (d≈-2.8). They also showed lower performance on grammar (d≈-1.5), phonology (d≈-0.9), mathematics (d≈-1.4), and on overall academic achievement (d≈-1.9). They were also 3.5 times as likely to have repeated a grade [14]. These results are partly due to our application of the “interference with academic performance” criterion. They may also partly be due to the fact that all those academic skills were tested in writing exclusively. Nevertheless, they are consistent with a large literature documenting weaknesses in many domains of oral language in dyslexia, but also in mathematics [26–30]. Although the two classifications largely agreed in this respect, it may be noted that mean scores in mathematics and overall academic achievement were slightly lower for pupils diagnosed with dyslexia by DSM-5 than by ICD-11, and their incidence of grade repetition was higher. This may be interpreted as a consequence of including more low-IQ children in the DSM-5 diagnosis.

#### Sociological variables

On average, pupils diagnosed with dyslexia had a lower SES (d≈-0.7), and they were twice as likely to be in a priority education (i.e., disadvantaged) area. This is consistent with a large literature on the impact of social factors on reading ability [13,31–33]. Interestingly, although it has been hypothesized that the IQ discrepancy criterion allowed one to better identify cases of dyslexia with a predominantly biological (rather than social) origin (e. g. [31]), in the present study the mean SES of the diagnosed population did not differ between the two classifications.

#### Sex ratio

The Programme for International Students Assessment (PISA) has now well documented sex differences in reading achievement: in France in 2015, boys’ reading performance was 29 points lower than girls’, with other OCDE countries showing the same trend (27 point mean difference) [34]. Consistently, many studies have reported that dyslexia affected boys more than girls. For instance, the Isle of Wight study [25] reported a sex ratio of about 3.3 boys to 1 girl for specific reading retardation. Since then, sex ratios have tended to decrease with successive studies (around 2 in [10,35]; 1.4 to 3 in [36]), reaching a low point close to 1 in [30] for poor reading (and 1.2 for poor spelling). Indeed, it has been noted since [25] that the sex-ratio was higher for specific reading disorder (involving a discrepancy criterion) than for poor reading (they reported 1.3). Our data are consistent with this observation: we find a sex ratio of 1.8 according to the ICD-11 definition, vs. 1.6 according to the DSM-5 or to a poor reading threshold. Furthermore, the more stringent the severity threshold, the larger the sex ratio (up to 2.2 at -2 SD). These results, based on a sample free from ascertainment bias, confirm that boys are at higher risk of reading disability, and particularly so of severe and specific reading disability.

#### Handedness

Historically, Geschwind and Behan [37] reported a significant association between handedness, dyslexia and other disorders. They proposed an explanatory model involving foetal testosterone that is still controversial, and replications of the findings have been mixed [38,39].. Another explanation could be pathological left-handedness, a condition linking some cases of left-handedness to intellectual retardation, due to birth trauma [40], but this is also controversial [41].

In the present study, we found a slight excess of non-right-handedness in dyslexia, which was significant using DSM-5 criteria but not using ICD-11. However, analyses adjusting on sex and IQ suggested that there is no specific association between dyslexia and non-right-handedness. Finally, our results are consistent with [23], in suggesting that high-IQ children are at much lower, not higher, risk of dyslexia.

### Limitations

The main limitations of this study have already been evoked and may be summarised as follows:

- Some diagnostic criteria could not be applied because the relevant information was not available in the database (see Table 1). These include: 1) “provision of interventions that target those difficulties” (DSM-5); 2) sensory impairment; 3) mental or neurological disorders. The first criterion would be particularly likely to modify prevalence rates and some of the comparisons between the two classifications if applied. However, we are not aware of any previous study applying it.
- Some diagnostic criteria were only approximated using available data. These include 1) lack or inadequacy of instruction, for which only serious illnesses were taken into account; 2) lack of proficiency in the language of academic instruction, for which only age of arrival in France was taken into account; 3) psychosocial adversity, for which only pupils in care were excluded.
- The only reading measure was reading comprehension, which may have inflated the reading-IQ correlation and the impact of IQ-based criteria, and underrepresented decoding-based poor readers. Furthermore, relying on only one reading measure made it less reliable than would be desirable, which would be a problem for individual diagnosis, but is less so for the present purposes.
- Reading and IQ tests were administered collectively in class, making them less reliable than individual evaluation by a trained professional, and perhaps inflating their correlation with school achievement, thus perhaps diminishing the impact of the achievement criterion.
- The results obtained are of course limited to the population of French 6^th^-graders in 2007, and are expected to vary depending on grade, language, and educational system.

Despite these limitations, this study is to our knowledge 1) the one using the largest subset of all DSM and ICD criteria; 2) the only one to systematically compare DSM-5 and ICD-11, including the differential effects of various criteria and thresholds; 3) one of those relying on the largest population samples, using weights to adjust the results to a representative sample of the population.

## Conclusion

We calculated the prevalence of dyslexia in France using a large database representative of French sixth-graders, and according to two different official definitions: the ICD-11’s and the DSM-5’s. Applying a -1.5 SD threshold, 3.5% and 6.6% of grade 6 students were identified as dyslexic, according to ICD-11 and DSM-5, respectively. However, the two definitions disagreed on the classification of a large number of students. Differences directly followed from the use by ICD-11 (but not DSM-5) of a discrepancy criterion between IQ and reading performance. Boys and students with lower SES and schooled in disadvantaged areas were overrepresented amongst those diagnosed with dyslexia, regardless of the definition. Furthermore, pupils with dyslexia also had lower scores in mathematics, non-verbal IQ, self-efficacy and motivation on average.

## Data Availability

Data can be requested from the ADISP archive of French public statistical data: http://www.progedo-adisp.fr/enquetes/XML/lil.php?lil=lil-0955. Under the terms of the data transfer agreement, we are not allowed to redistribute it.

## Declarations

### Ethics approval

The study was approved by the National Council for Statistical Information (CNIS) (visa n°2008A061ED and 2011A082ED), ensuring public interest and conformity with ethical, statistical and confidentiality standards.

### Consent to participate

Participation was compulsory as part of the evaluation policy of the French Ministry of National Education.

### Consent for publication

Not applicable.

### Code availability

Analysis scripts are available on: https://osf.io/kejcp/?view_only=c0736a7a007346218ef1578f8d760e8c

### Authors’ contributions

FR designed the study, CDF and AG performed the analysis under supervision of HP and FR, CDF and FR wrote the paper, all authors revised the paper and approved the final version.

## Acknowledgements

The data used was obtained from « Panel d’élèves du second degré, recrutement 2007 - 2007-2013, DEPP - Ministère de l’Éducation [producteur], ADISP-CMH [diffuseur] ».

