## Supplementary material for "Epidemiology of developmental dyslexia: A comparison of DSM-5 and ICD-11 criteria"

### Supplementary Information for : Epidemiology of developmental dyslexia: A comparison of DSM-5 and ICD-11 criteria

European Journal of Epidemiology

### Supplementary Methods

**Figure S1.** Flowchart of Included and Excluded Participants

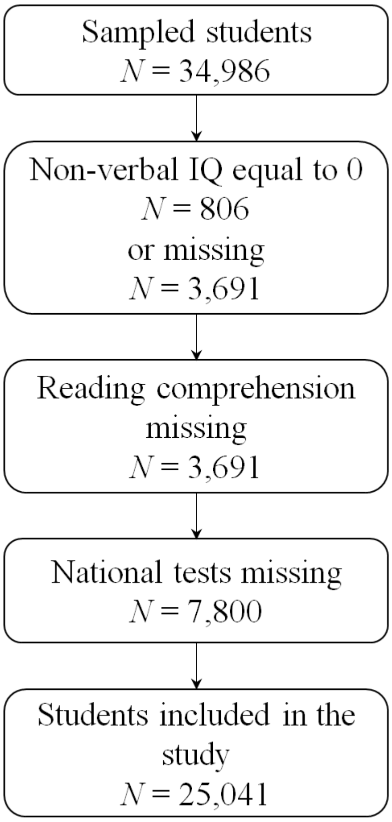

Table S1. Descriptive statistics for excluded and included participants. Source : MENESR DEPP, Panel 2007.

| Outcome variable | **Excluded N=9,945** | | **Included N=25,041** | | **Difference (Excluded-Included)** | |
|---|---|---|---|---|---|---|
| | *M (or %)* | *SD* | *M (or %)* | *SD* | *d or Odds Ratio* | *p* |
| *Reading Scores* | | | | | | |
| Reading comprehension (15 items) | 8.72 | 3.66 | 9.63 | 3.08 | -0.27 | <.0001 |
| Grammar (20 items) | 7.69 | 4.64 | 8.85 | 4.31 | -0.26 | <.0001 |
| Phonology (10 items) | 6.29 | 2.46 | 6.83 | 2.16 | -0.23 | <.0001 |
| Mathematics (48 items) | 23.94 | 10.02 | 26.63 | 8.77 | -0.29 | <.0001 |
| Intelligence score (non-verbal) (30 items) | 13.12 | 7.64 | 15.75 | 5.94 | -0.38 | <.0001 |
| National assessment score (out of 100) | 51.58 | 19.78 | 60.47 | 17.11 | -0.48 | <.0001 |
| *Perceived self-efficacy* | | | | | | |
| Self-regulation (z-score) | -0.05 | 1.03 | 0.04 | 0.97 | -0.09 | <.0001 |
| Social self-efficacy (z-score) | -0.05 | 1.05 | 0.02 | 0.97 | -0.07 | <.0001 |
| Academic self-efficacy (z-score) | -0.08 | 1.06 | 0.02 | 0.98 | -0.10 | <.0001 |
| *Motivation* | | | | | | |
| Intrinsic motivation (z-score) | 0.00 | 0.93 | -0.01 | 0.92 | 0.01 | 0.5299 |
| Extrinsic motivation (z-score) | 0.00 | 0.84 | -0.01 | 0.82 | 0.02 | 0.2618 |
| Amotivation (z-score) | 0.05 | 0.94 | -0.02 | 0.87 | 0.07 | <.0001 |
| Socio-economic index (z-score) | -0.14 | 1.07 | 0.05 | 0.97 | -0.18 | <.0001 |
| Priority Education (%, OR) | 19.9 | | 13.4 | | 1.61 | <.0001 |
| Boys (%, OR) | 52.9 | | 50.4 | | 1.11 | <.0001 |
| Left-handed (%, OR) | 11.0 | | 11.8 | | 0.99 | 0.7404 |
| Repeater (%, OR) | 25.5 | | 15.7 | | 4.52 | <.0001 |

#### Measures – examples of test items

##### Reading comprehension

Example: D'après le texte, combien de garçons escaladent le mur ?...

(According to the text, how many boys are climbing on the wall?.…)

##### Phonological awareness

Example: fer, aimer, verre, amer, hiver (\fɛʁ\, \e.me\, \vɛʁ\, \a.mɛʁ\, \i.vɛʁ\)

##### *Grammar*

Example: « Septembre ! C'est le mois.....choisit l'hirondelle pour partir vers le sud du Sahara.....elle peut passer l'hiver au chaud ». Correct answers : « que », « où ».

("September! This is the month.....the swallow chooses to fly towards the South of Sahara....it can spend its winter warm"). Correct answers: "that", "where".

##### Mathematics

Examples:

- 27 × 20 = ...

- Zoé est plus petite que Joëlle, et elle est plus grande que Cécile. La fille la plus grande s'appelle : 1) Cécile 2) Zoé 3) Joëlle (Zoé is shorter than Joëlle, and she is taller than Cécile. The tallest girl is called: 1) Cécile 2) Zoé 3) Joëlle)

#### Weighted least squares

For the ICD-11 definition, we estimated the variance of the residuals of reading comprehension on non-verbal intelligence score as a function of non-verbal intelligence scores. We first regressed the squared residuals of the OLS regression on intelligence score. The fitted values from this regression constitute our estimate of residual variance. We then ran weighted least squares using the inverse of this estimate as weights in order to produce efficient regression parameters.

#### Statistical analyses

##### Non-response propensity weights

In order to account for differences in the likelihood to respond to the survey, we computed non-response propensity weights, which we combined with the initial weights from the exhaustive baseline survey. Non-response propensity scores were first computed with a logistic regression, and inversed in order to obtain the non-response propensity weights. Both the baseline weights and the non-response propensity weights were then scaled; the product of these two scaled weights constitutes our final weights. The R code can be found below:

#### Supplementary results

Table S2. Descriptive statistics in the working sample, computed with non-response propensity weights. Source : MENESR DEPP, Panel 2007.

| Variables | N | Mean or % | SD | Min | Max |
|---|---|---|---|---|---|
| *Reading Scores* | | | | | |
| Reading comprehension | 25041 | 9.47 | 3.12 | 0.00 | 15.00 |
| Grammar | 25041 | 8.61 | 4.32 | 9.00 | 20.00 |
| Phonology | 25041 | 6.74 | 2.19 | 0.00 | 10.00 |
| Mathematics | 25041 | 25.98 | 8.82 | 0.00 | 48.00 |
| National assessment average score | 25041 | 59.33 | 17.35 | 0.00 | 98.50 |
| Intelligence score (non-verbal) | 25041 | 100.00 | 15.00 | 62.80 | 136.41 |
| *Perceived self-efficacy* | | | | | |
| Self-regulation | 23826 | 0.01 | 0.99 | -2.24 | 0.88 |
| Social self-efficacy | 22644 | 0.00 | 0.99 | -6.11 | 1.66 |
| Academic self-efficacy | 21709 | 0.00 | 0.99 | -5.30 | 2.12 |
| *Motivation* | | | | | |
| Intrinsic motivation | 23291 | -0.01 | 0.92 | -3.02 | 1.38 |
| Extrinsic motivation | 23291 | -0.01 | 0.83 | -2.14 | 1.72 |

| | | | | | |
|---|---|---|---|---|---|
| Amotivation | 23291 | 0.00 | 0.88 | -0.79 | 4.43 |
| Socio-economic index | 23536 | 0.00 | 1.00 | -5.92 | 4.31 |
| Priority Education (%) | 25041 | 15.6 | | | |
| Boys (%) | 25041 | 51.0 | | | |
| Left-handed (%) | 23727 | 11.9 | | | |
| Repeater (%) | 25041 | 17.1 | | | |

Table S3. Sex ratio (boys/girls) as a function of diagnostic criteria. Source : MENESR DEPP, Panel 2007.

| | Entire popu-lation | Reading < threshold | Reading < threshold + one exclusion criterion | | | | Reading < threshold + all exclusion criteria | Reading < threshold + Academic achievement < -0.5 SD | All preceding criteria (DSM-5) | All preceding criteria + reading-IQ discrepancy (ICD-11) |
|---|---|---|---|---|---|---|---|---|---|---|
| Severity threshold | | | Serious illness | Language proficiency | Psychosocial adversity | IQ<70 | | | | |
| -1 SD | | 1.47 | 1.46 | 1.47 | 1.47 | 1.42 | 1.41 | 1.49 | 1.41 | 1.53 |
| -1.25 SD | | 1.58 | 1.54 | 1.57 | 1.56 | 1.51 | 1.5 | 1.68 | 1.51 | 1.57 |
| **-1.5 SD** | 1.04 | 1.61 | 1.61 | 1.61 | 1.61 | 1.54 | 1.54 | 1.65 | 1.58 | 1.83 |
| -1.75 SD | | 1.76 | 1.75 | 1.78 | 1.77 | 1.69 | 1.7 | 1.78 | 1.73 | 2.11 |
| -2 SD | | 1.76 | 1.75 | 1.78 | 1.77 | 1.69 | 1.7 | 1.78 | 1.73 | 2.22 |

Handedness and dyslexia

Table S4. Prevalence of non-right-handedness as a function of diagnostic criteria. Source : MENESR DEPP, Panel 2007.

| | Entire population | Reading < threshold | Reading < threshold + one exclusion criterion | | | | Reading < threshold + all exclusion criteria | Reading < threshold + Academic achievement < -0.5 SD | All preceding criteria (DSM-5) | All preceding criteria + reading-IQ discrepancy (ICD-11) |
|---|---|---|---|---|---|---|---|---|---|---|
| Severity threshold | | | Serious illness | Language proficiency | Psychosocial adversity | IQ<70 | | | | |
| -1 SD | | 15,0 [a] | 15,0 | 15,3 | 15,0 | 15,1 | 15,3 | 15,0 | 15,3 | 15.2 n.s. |
| -1.25 SD | | 15,2 | 15,3 | 15,5 | 15,3 | 15,2 | 15,6 | 15.0 n.s. | 15,4 | 15.4 n.s. |
| **-1.5 SD** | 13,8 | 15,7 | 15,8 | 16,0 | 15,8 | 15,9 | 16,5 | 15,5 | 16,4 | 14.9 n.s. |
| -1.75 SD | | 16,5 | 16,7 | 17,0 | 16,6 | 16,8 | 17,9 | 15.4 n.s. | 16,7 | 14.7 n.s. |
| -2 SD | | 16,5 | 16,7 | 17,0 | 16,6 | 16,8 | 17,9 | 15.3 n.s. | 16,7 | 13.4 n.s. |

[a] All prevalence rates in columns 2-10 are significantly different from that in the entire population (Column 1) at $p<0.05$, except where indicated as n.s..

Logistic regression with dyslexia as dependent variable and non-righthandedness, sex and IQ as independent variables:

With DSM-5 definition:

| | Estimate | Std. Error | z value | Pr(>\|z\|) |
|---|---|---|---|---|
| (Intercept) | 2,924 | 0,304 | 9,631 | 0,000 |
| sex | -0,879 | 0,387 | -2,273 | 0,023 |
| IQ | -0,062 | 0,003 | -18,745 | 0,000 |
| handedness | 0,539 | 0,775 | 0,695 | 0,487 |
| sex:IQ | 0,014 | 0,004 | 3,308 | 0,001 |
| sex:handedness | 0,148 | 0,998 | 0,149 | 0,882 |
| IQ:handedness | -0,002 | 0,009 | -0,219 | 0,827 |
| sex:IQ:handedness | -0,007 | 0,011 | -0,596 | 0,551 |

With ICD-11 definition:

| | Estimate | Std. Error | z value | Pr(>\|z\|) |
|---|---|---|---|---|
| (Intercept) | -0.62 | 0.43 | -1.46 | 0.14 |
| sex | -0.47 | 0.53 | -0.88 | 0.38 |
| IQ | -0.03 | <0.01 | -7.25 | <0.01 |
| handedness | 0.42 | 1.07 | 0.39 | 0.7 |
| sex:IQ | 0.01 | 0.01 | 1.98 | 0.05 |
| sex:handedness | -0.25 | 1.39 | -0.18 | 0.86 |
| IQ:handedness | <0.01 | 0.01 | -0.08 | 0.94 |
| sex:IQ:handedness | <0.01 | 0.01 | -0.27 | 0.79 |
